# Measurement Instruments Assessing Organizational Resilience in Health Facilities: A Systematic Review of Psychometric Properties

**DOI:** 10.1101/2025.06.11.25329057

**Authors:** Atena Pasha, Kaylin Morris, Karolina Kaczmarski, Xiaoming Li, Shan Qiao

## Abstract

**Background:** The COVID-19 pandemic has underscored the critical importance of resilient health systems. Organizational resilience, as a multidimensional construct encompassing cognitive, behavioral, and contextual capacities, is essential for ensuring continuity of care during crises. However, the availability of psychometrically sound instruments for assessing organizational resilience within healthcare facilities remains limited.

**Methods:** Following PRISMA guidelines, a comprehensive systematic review was conducted across PubMed, Embase, PsycINFO, CINAHL, and Web of Science to identify empirical studies conducted between January 2000 and May 2024 evaluating the psychometric properties of organizational resilience instruments applied in health systems. Titles, abstracts, and full texts were screened independently by two reviewers. Data extraction covered instrument characteristics, psychometric properties, and study methodology. The (Consensus-based Standards for the selection of health status Measurement Instruments) COSMIN checklist was used to assess methodological quality, and a standardized scoring system (1–10 scale) was applied across six core psychometric properties. Instruments were synthesized and compared based on measurement level (team and system), contextual scope, and methodological rigor. The review was pre-registered in PROSPERO (ID: CRD42024511040).

**Results:** Of the 7,479 records screened, 12 studies were included evaluating team-level (n = 7) and system-level (n = 5) instruments assessing organizational resilience in healthcare. Internal consistency was reported in eight studies, test-retest reliability in two studies, content validity in 10 studies, structural validity in 10 studies, criterion validity in no studies, and construct validity in all studies. At the team level, the Resilience Engineering questionnaire and at the system level, the Organizational resilience for public health centers emerged as the most psychometrically robust tools.

**Conclusions:** This review provides a critical synthesis of the available instruments for measuring organizational resilience in health facilities, revealing significant psychometric and methodological gaps. There is an urgent need to develop and validate context-specific, system-level resilience instruments incorporating intersectionality and cultural sensitivity.

## Introduction

The COVID-19 pandemic has profoundly hit the global healthcare system with significant challenges and disruptions in healthcare delivery. Health organizations have had to quickly adapt by reallocating and reorganizing resources to meet the needs of COVID-19 patients, amidst high demand for urgent healthcare and sustained periods of increased activity (1–3). During the pandemic, health facilities focused on adapting public health emergencies to provide patient care, address employee health concerns, and support employee psychological safety and mental health. However, many health facilities have struggled to demonstrate sufficient resilience to effectively cope with the crisis (4).

Organizational resilience refers to an organization’s ability to maintain functions and recover swiftly from adversity by mobilizing the necessary resources (5). In the context of health facilities, it encompasses how hospitals and healthcare clinics monitor and plan for fluctuations in demand, handle shocks or stressors, and adapt and learn (6). Improving the organizational resilience of health systems can reduce vulnerability to crises, better preparing them to respond effectively while minimizing disruption to essential healthcare services (4, 7). Policymakers and researchers increasingly rely on academic literature to comprehend how to build resilience in healthcare organizations and measure this concept (8). Consequently, there is a growing need to develop methods or metrics to measure and monitor organizational resilience in healthcare.

Despite growing interest, there remains no universally accepted definition or conceptual framework for organizational resilience in healthcare. Additionally, the number of studies focusing on organizational resilience is still small in terms of establishing a valid measurement scale for organizational resilience, with even fewer studies on developing resilience measurement frameworks and indices for the healthcare context. Measuring organizational resilience in healthcare facilities poses challenges due to the scarcity of validated indicators and measurement tools. Several models and metrics have been developed, but no universally accepted approach exists for measuring or assessing organizational resilience. As noted by Ignatowicz et al. (8), this ambiguity affects what is measured, how it is measured, and for what purpose.

The existing research has mostly focused on specific aspects of organizational resilience in relation to other variables instead of looking at organizational resilience as a whole (6). They either measure a set of characteristics such as knowledge, learning, planning, agility, adaptability, and robustness (9, 10) or evaluate capacity to deal with shocks and stresses (11, 12). However, their applicability to a healthcare context may be limited. This conceptual variation has also resulted in diverse assessment approaches in terms of scope, format, and intended use. It is imperative to note that existing reviews have not yet comprehensively evaluated the precise psychometric properties of the instruments encompassing both organizational resilience and healthcare facilities.

It is also noteworthy that, except for one review (8), none of the other reviews narrowed their inclusion criteria to studies that exclusively assessed at least one psychometric property. Although Ignatowicz et al. (8) offered an important synthesis of resilience assessment approaches in healthcare, their review did not systematically appraise the psychometric rigor or methodological quality of the identified instruments. Therefore, it is crucial to develop robust and conceptually sound approaches for assessing organizational resilience in order to ensure that the measures are reliable, robust, and truly reflect the realities faced by healthcare facilities.

The psychometric properties of reliability and validity are fundamental aspects that need to be considered when assessing the effectiveness of an instrument (13). Reliability measures the consistency of results over time and space, and there are several metrics used to assess reliability, including Cronbach’s alpha and test-retest reliability (13, 14). Cronbach’s alpha assesses the internal consistency of the scale (15), while test-retest reliability evaluates the stability of the instrument’s results over time (13). On the other hand, validity addresses whether the instrument accurately measures the intended construct (16). It is essential to consider various types of validity, such as content validity, structural validity, construct validity, and criterion validity.

Therefore, the aim of this study was to bridge the knowledge gap and enhance our understanding of the psychometric properties of measurement instruments for organizational resilience in health facilities. Through a comprehensive systematic review, we evaluated the psychometric properties of various instruments and identified the most robust options. In this review, organizational resilience was defined as the capacity of health facilities or systems to withstand, adapt to, and recover from disruptions while continuing to deliver essential health services. Given the common use of team self-reports to reflect organizational resilience within health facilities, this review also explored team-based measures for organizational resilience. The methodological framework was primarily informed by the measurement properties outlined by Terwee et al. (17). Their comprehensive approach provided quality criteria for various psychometric properties, including content validity, internal consistency, criterion validity, construct validity, reproducibility (test-retest reliability), structural validity, responsiveness, and interpretability. We focused on evaluating all except responsiveness and interpretability. Responsiveness detects clinically important changes over time (18). Interpretability identifies whether the score difference between groups is meaningful (19). Our emphasis was more on establishing the fundamental measurement properties, such as reliability and validity, at a single point in time for a single group to ensure a robust foundation for any future psychometric assessments with longitudinal data and multiple groups (13).

## Materials and Methods

### Search Strategies and Data Sources

The protocol for this systematic review adhered to the PRISMA (Preferred Reporting Items for Systematic Reviews and Meta-Analyses) guidelines (20). The search focused on studies published from January 1, 2000, to May 2024. This time frame was chosen to include measurement tools developed or utilized in the past two decades, a period marked by increasing interest in organizational resilience frameworks in response to significant health system shocks, such as SARS, Ebola, and COVID-19. The review was pre-registered in PROSPERO (ID: CRD42024511040).

The authors, KM and KK, carried out a comprehensive search from March to May 2024 across five bibliographic databases: PubMed, Embase, PsycINFO, CINAHL, and Web of Science. The reference lists of reviews and meta-analyses, as well as the reference lists of included studies, were also examined (see **Appendix 1**). We excluded grey literature (e.g., WHO, USAID reports) and JBI from our current review, given our focus on psychometrically validated, peer-reviewed tools. However, we acknowledge that this exclusion may limit the identification of unpublished tools used in field settings. To ensure comprehensiveness, we cross-checked our search approach and included studies against recent reviews (8, 21).

The search strategy included keywords from three categories. The first category comprised keywords related to health facilities, such as “Health Facilities,” “Health Services,” and “Health Delivery.” Then, to ensure the focus remained on organizational- or system-level resilience constructs, search terms were designed to include both broad resilience constructs (e.g., “resilien”) and organization-specific modifiers (e.g., “organizational resilien”). Studies solely measuring individual or trait-based personal resilience were excluded during screening. The third category included keywords related to measurement, such as “Measure,” “Surveys,” and “Questionnaires.”

Keywords were organized using the following logical operators: (1) keywords within one category were linked using the OR operator (e.g., Healthcare worker OR Health System), and (2) keywords across different categories were connected using the AND operator (e.g., Health Service AND organizational resilience AND measure) (see **Appendix 1**).

### Eligibility Criteria

The criteria for inclusion in this study were two-fold. First, the article must be published in English in a peer-reviewed journal, and second, the study must evaluate at least one psychometric property of an instrument that measures the organizational resilience of health facilities or team-based reports in any type of measures (self-reported, expert-rated, or expert panel reviews). Eligible studies employed structured measurement instruments to assess resilience at team or system levels. Excluded papers were those that claimed to measure organizational resilience but did not utilize a corresponding resilience scale, as well as studies that measured only individual-level resilience. Additionally, papers not published in English were excluded from the review.

To further distinguish between individual- and organizational-level resilience, we adopted an operational definition of organizational resilience as “the capacity of health facilities or systems to withstand, adapt to, and recover from disruptions while continuing to deliver essential health services.” Instruments were included only if the resilience construct referred to organizational, team, or system-level functioning, rather than individual psychological traits or behaviours.

Specifically, we excluded tools measuring individual or staff-level resilience unless the unit of analysis and scale design clearly targeted organizational processes, structures, or collective performance (e.g., coordination, leadership, system learning, or resource mobilization). To increase conceptual clarity, the screening process also drew on theoretical models (e.g., resilience engineering, socio-ecological resilience) and alignment with prior frameworks used by Ignatowicz et al. (8). In cases of ambiguity, three reviewers discussed the theoretical underpinning of the instrument and reached consensus on inclusion.

Included studies were screened based on the eligibility criteria using the data management software RAYYAN (22). The study selection process consisted of three phases. First, all the duplicate studies were removed. Second, researchers (AP, KM, KK) screened all titles and abstracts. Studies that did not comprise an instrument assessing organizational resilience among health facilities were removed. Third, two researchers (KM, AP) conducted full-text screening of the remaining articles and removed studies that considered resilience only at the individual level, did not assess at least one psychometric property, or collected only qualitative data. Finally, articles that met the inclusion criteria were selected for this review (see **Fig 1**). Any discrepancies were resolved through discussion (KM, AP). In cases where the agreement could not be reached, a third researcher (SQ) was involved to adjudicate.

**Fig 1.** Flow diagram of the literature search and articles selection (adapted from PRISMA 2020 guidelines for systematic reviews) (23).

### Data Extraction

Key information on three main categories was extracted from each included study, including study features, psychometric properties of the instrument, and the methodological quality of the study. In terms of study features, data were extracted based on the name of the instrument developed and/or validated, the description of the instrument (factors, items, and response scale), population, sample size, and country of data collection. As for the psychometric properties of the instrument, data were extracted on internal consistency, test-retest reliability, content validity, structural validity, criterion validity, and construct validity. Information on study design and methodology was evaluated and rated for methodological quality assessment. Any discrepancies among data extractors (AP, KM) were resolved through group discussions. In cases where the agreement could not be reached, a third researcher (SQ) was involved to adjudicate. This multi-stage review process ensured that all eligible studies were systematically identified and included in the final analysis.

### Psychometric Properties Assessment

The Consensus-based Standards for the selection of health status Measurement Instruments (COSMIN) checklist (24) comprises nine boxes, each evaluating the methodological quality of a specific psychometric property. The number of items in these boxes ranges from 5 to 18. Each item within a box is rated as “yes”, “not applicable (NA)”, or “not reported (NR)”. One item, “Were there any important flaws in the methods or design of the study?” has been removed due to subjectivity. Additionally, the item, “Was the sample size adequate?”, has been considered as well as whether an appropriate analysis was conducted to identify an adequate sample size, such as a power analysis.

Psychometric properties assessment was evaluated using selected boxes from the COSMIN checklist. For each psychometric property (internal consistency, test–retest reliability, content validity, structural validity, criterion validity, and construct validity), COSMIN items were rated strictly based on what was explicitly reported in each article (see **Appendix 2**). Items were scored as 1 (“Yes”) if adequately reported and 0 (“Not reported”); items rated as “Not applicable” were excluded from scoring. The COSMIN item addressing general design flaws was removed, and sample size items were scored as “Yes” only when an a priori sample size justification was reported. For each property, the mean item score was calculated and rescaled to a 0–10 scale. Scores of 7.1–10 indicated strong, 3.1–7 moderate, and 1–3 limited methodological quality, with a score of 0 indicating no evidence.

### Methodological Quality Assessment

The methodological quality of the studies included in this review was assessed using the COSMIN checklist. COSMIN is a widely recognized tool for evaluating the quality of studies that examine the psychometric properties of patient-reported outcome measures (PROMs). It is important to note that although COSMIN was developed for PROMs, it remains the most comprehensive and widely accepted framework for evaluating measurement quality. However, its application to instruments that assess organizational or system-level resilience, such as those focused on healthcare facilities, requires careful adaptation.

Given that no validated alternative appraisal tool exists for organizational or system-level resilience measures, COSMIN offers a transparent, reproducible, and theoretically grounded method for assessing measurement rigor in this review. COSMIN is based on Classical Test Theory, which considers observed scores as a combination of a true score and random error, providing a systematic framework for evaluating key psychometric properties of measurement instruments (25). Its structured, domain-based approach, which covers reliability, validity, and responsiveness, can be effectively applied to organizational instruments when interpreted correctly.

To the best of our knowledge, currently there is no established methodological quality checklist specifically designed for organizational or system-level resilience measures in healthcare settings. As a result, we applied the COSMIN checklist to all included studies, regardless of whether the measurement level was team or system. When necessary, we interpreted COSMIN domains within the organizational context, especially for aspects such as content validity, structural validity, and reliability, ensuring their relevance to system-focused instruments. This pragmatic approach aligns with previous psychometric reviews in emerging fields, where existing methodological frameworks have been thoughtfully adapted due to the lack of domain-specific appraisal tools (26).

The assessment of psychometric properties and methodological quality was conducted by two independent reviewers (AP, KM). Any discrepancies in ratings were discussed and resolved among the reviewers. In cases where the agreement could not be reached, a third researcher (SQ) was involved to adjudicate. The involvement of multiple independent reviewers and resolving discrepancies likely enhances the reliability and consistency of the assessment process using the COSMIN checklist.

### Data Synthesis

An additional scoring framework was created to enable comparisons among all instruments with respect to their psychometric and methodological quality (24) (see **Appendix 3**). This framework assigned numerical values to each evaluated property based on its psychometric rating and methodological rigor. Properties receiving a “+” psychometric rating were assigned scores of 3, 2, or 1 when supported by strong, moderate, or limited methodological quality, respectively. Conversely, properties with a “−” psychometric rating were assigned scores of −1, −2, or −3 for strong, moderate, or limited methodological quality, respectively. Higher total scores reflected superior psychometric and methodological quality. Scores across all psychometric properties were subsequently summed to generate an overall score for each instrument, allowing for a quantitative comparison of instruments based on their combined psychometric and methodological performance.

## Results

### Characteristics of the Reviewed Studies

The characteristics of the reviewed studies are summarized in **Table 1**. Of the 7,479 records initially identified [MEDLINE (PubMed) (n = 1603), Web of Science (Clarivate) (n = 1976), PsycINFO (EBSCOhost) (n = 760), CINAHL (EBSCOhost) (n = 891), Embase (Elsevier) (n = 2249)], 12 studies were ultimately included in this systematic review. These studies evaluated the psychometric properties of instruments designed to measure resilience within health facility contexts. While our review focused on instruments assessing organizational resilience, we also identified and included instruments assessing resilience at the team level due to their use in health facility settings.

**Table 1.**
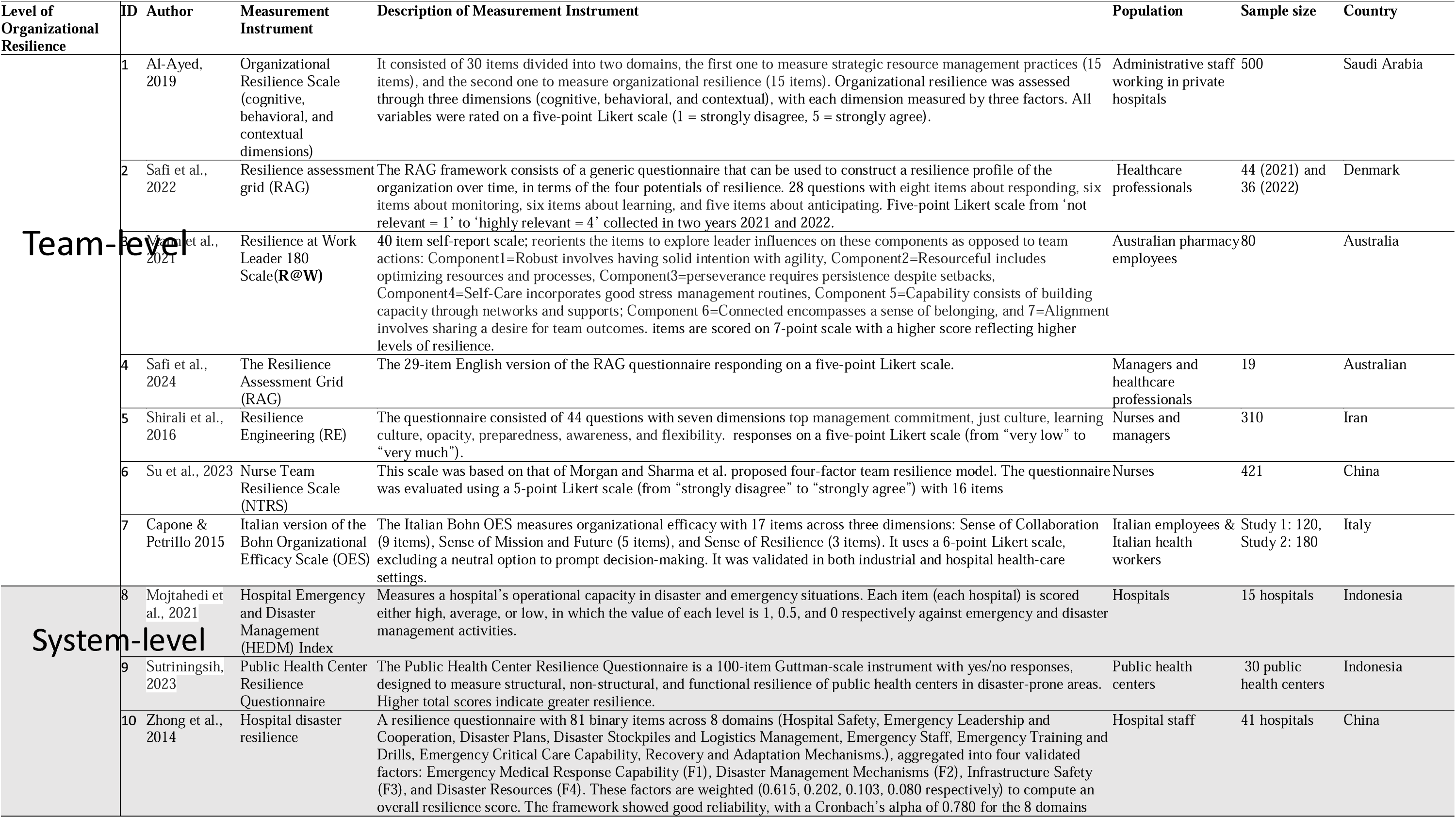
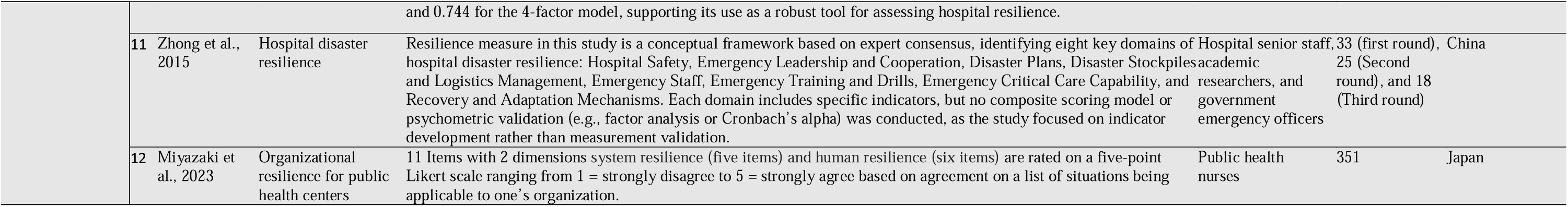
Characteristics of Included Studies.

The instruments included in this review reflect diverse conceptualizations of resilience, spanning team capacities (e.g., adaptability and leadership) (27–32), and organizational capabilities (e.g., preparedness, monitoring, learning, risk management) (33–38). At the team level, resilience was conceptualized as a behavioral factor enabling teams to adapt and recover from workplace stress. At the system level, resilience referred to the institutional capacity to maintain functionality, adapt during disruptions, and ensure continuity of care.

The reviewed instruments encompassed a variety of constructs, such as team cohesion, leadership adaptability, culture, system preparedness, and organizational learning. For instance, the Nurse Team Resilience Scale (NTRS) (31) emphasized intra-team dynamics, while the Resilience Assessment Grid (RAG) (30) and the Resilience Engineering (37) focused on institutional and systemic resilience performance.

Studies were geographically diverse, spanning countries such as China (31, 35, 36), Indonesia (33, 34), Australia (28, 32), Iran (37), Japan (38), Denmark (30), Italy (29), and Saudi Arabia (27). Participants ranged from frontline healthcare workers and nurses to administrators and system-level decision-makers. At the team level, sample sizes varied widely from studies with 19 hospital staff (36) to larger-scale studies involving over 500 respondents (27). Regarding system level, there were studies among 15 (35) to 41 hospitals (34).

Studies examining resilience in healthcare contexts have primarily utilized self-reported questionnaires, often employing Likert scales and psychometric validation techniques such as exploratory and confirmatory factor analyses (EFA and CFA) to assess constructs including organizational and team resilience. For example, Al-Ayed (27) developed a 30-item self-reported questionnaire to measure strategic human resource management practices and organizational resilience dimensions (cognitive, behavioral, contextual), distributed to hospital staff, and analyzed via EFA and CFA. Similarly, Su et al. (31) validated an 8-item NTRS using Likert-scale self-reported responses from nurses. The RAG was adapted and applied via Likert-scale questionnaires in internal medicine and outpatient settings, with content validation through modified Delphi methods and radar chart analyses (30, 32). Other studies incorporated the Delphi consensus for item development (36) or TOPSIS multi-attribute decision-making for indexing hospital resiliency (33), while factor analysis underpinned framework validation in disaster resilience self-reported assessments (34, 35, 37).

### Psychometric Properties and Their Methodological Quality of Instruments

A summary of the psychometric properties of instruments is presented in **Table 2**, and a summary of the methodological quality assessment is presented in **Table 3**. Based on the scoring system (**Appendix 2 and 3**), the synthesized psychometric and methodological quality assessment results are presented in **Table 4**.

**Table 2.**
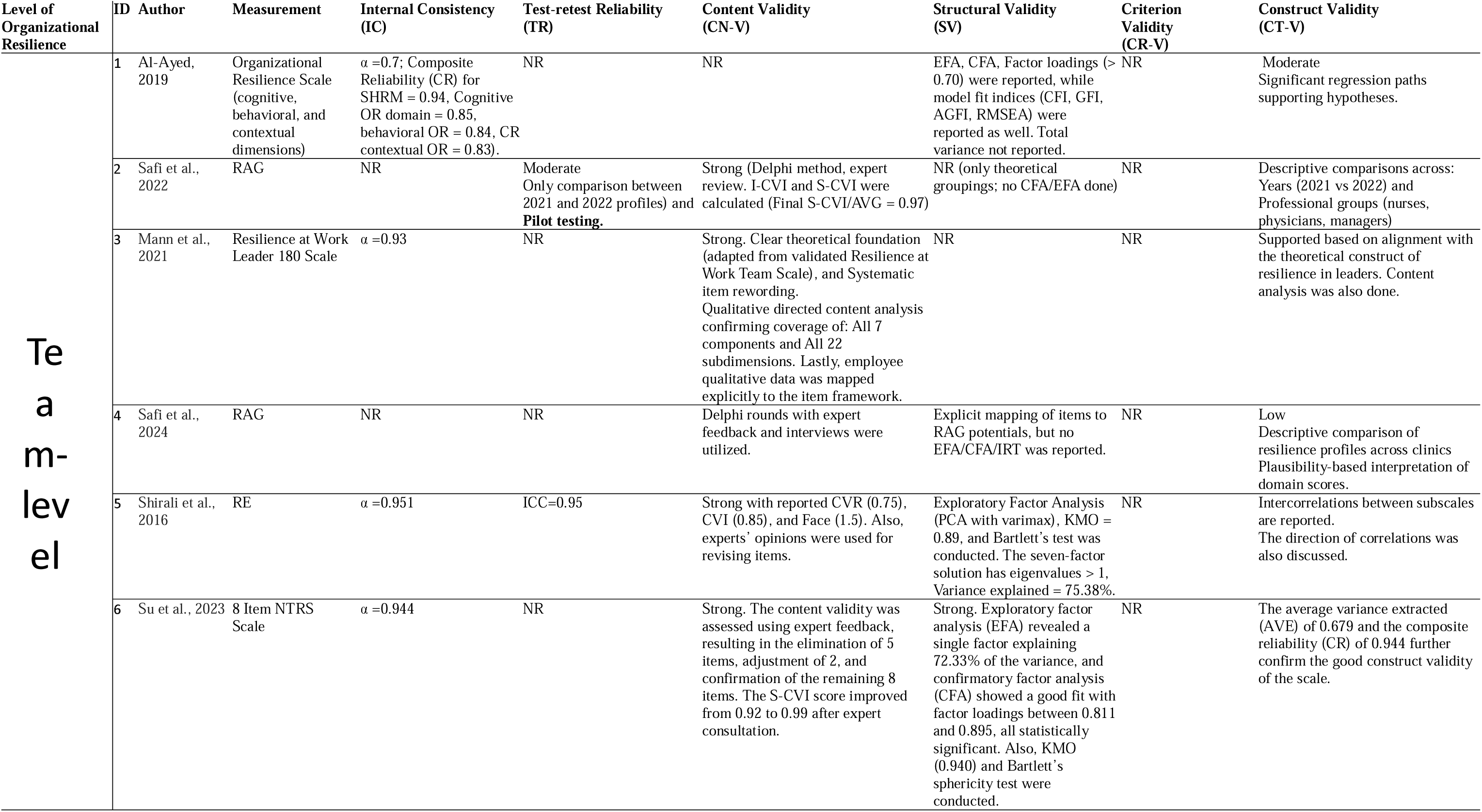
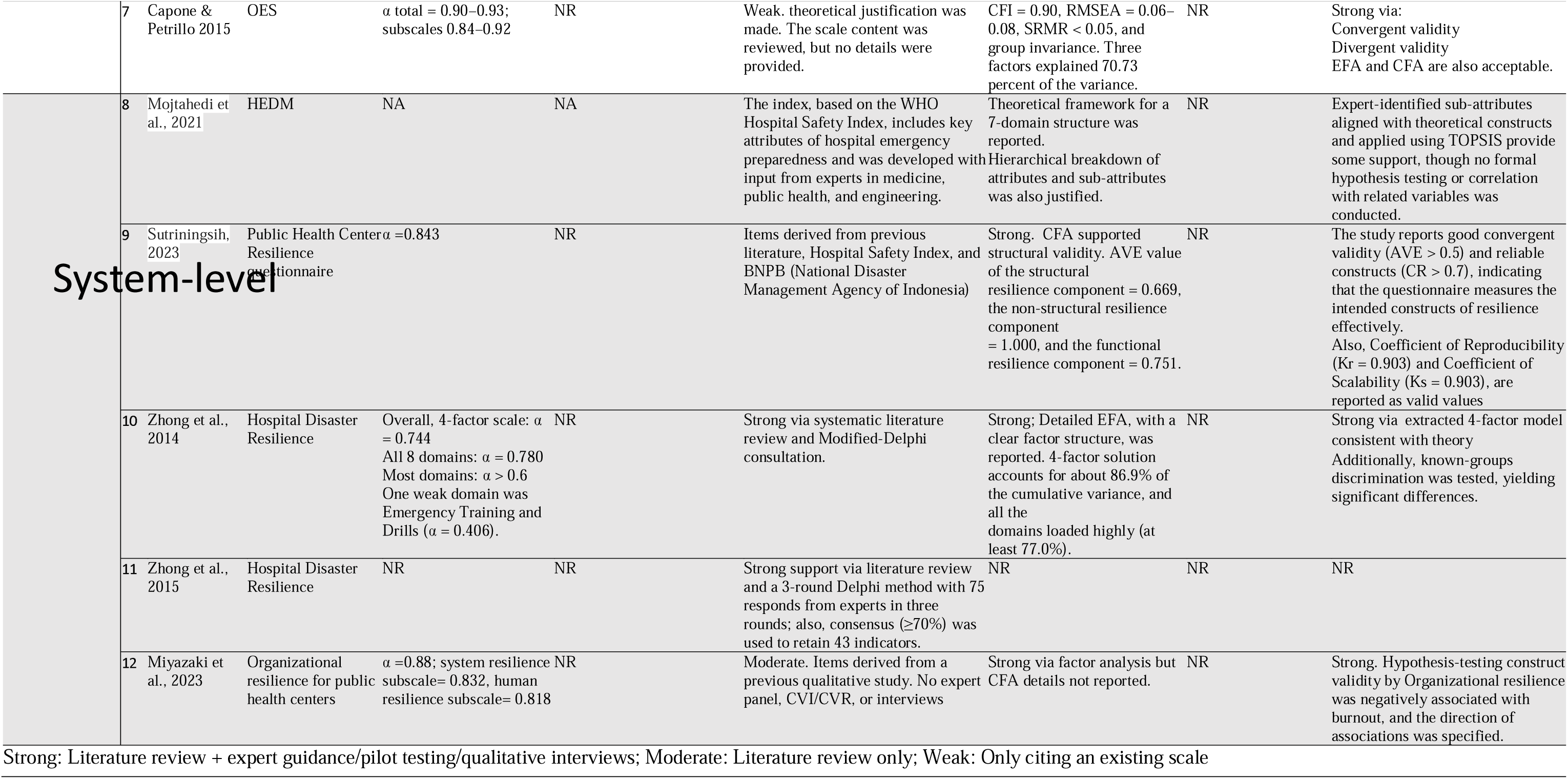
Summary of psychometric properties.

**Table 3.** Methodological quality assessment for the psychometric properties.

| Level of Organizational Resilience | ID | Author | Tool | Internal Consistency (IC) | Test-retest Reliability (TR) | Content Validity (CN-V) | Structural Validity (SV) | Criterion Validity (CR-V) | Construct Validity (CT-V) |
| --- | --- | --- | --- | --- | --- | --- | --- | --- | --- |
| Team-level | 1 | Al-Ayed, 2019 | Organizational Resilience Scale (cognitive, behavioral, and contextual dimensions) | 6.25 | NR | NR | 6 | NR | 5.71 |
|  | 2 | Safi et al., 2022 | RAG | NR | 2 | 10 | 2 | NR | 1.5 |
|  | 3 | Mann et al., 2021 | Resilience at Work Leader 180 Scale | 5.5 | NR | 8.5 | 2 | NR | 7.5 |
|  | 4 | Safi et al., 2024 | RAG | NR | NR | 10 | 1 | NA | 1 |
|  | 5 | Shirali et al., 2016 | RE | 9 | 5 | 10 | 9 | NA | 4 |
|  | 6 | Su et al., 2023 | NTRS | 8 | NR | 10 | 8 | NR | 7 |
|  | 7 | Capone & Petrillo, 2015 | OES | 8 | NR | NR | 8 | NR | 10 |
| System-level | 8 | Mojtahedi et al., 2021 | HEDM | NA | NA | 8 | 1 | NA | 1 |
|  | 9 | Sutriningsih, 2023 | Public Health Center Resilience questionnaire | 6 | NA | 8 | 7 | NA | 5 |
|  | 10 | Zhong et al., 2014 | Hospital Disaster Resilience | 6 | NA | 7.5 | 8 | NR | 7.1 |
|  | 11 | Zhong et al., 2015 | Hospital Disaster Resilience | NR | NR | 9 | NR | NR | NR |
|  | 12 | Miyazaki et al., 2023 | Organizational resilience for public health centers | 9.5 | NA | 4 | 9 | NR | 7.1 |

\*Scores transformed to a scale of 1-10
Methodological Quality (MQ) ratings: On a scale of 1-10, a methodological rating of 1-3 was “limited,” 3.1-7 was “moderate,” and 7.1-10 was “strong” (see electronic supplementary material).

**Table 4.**
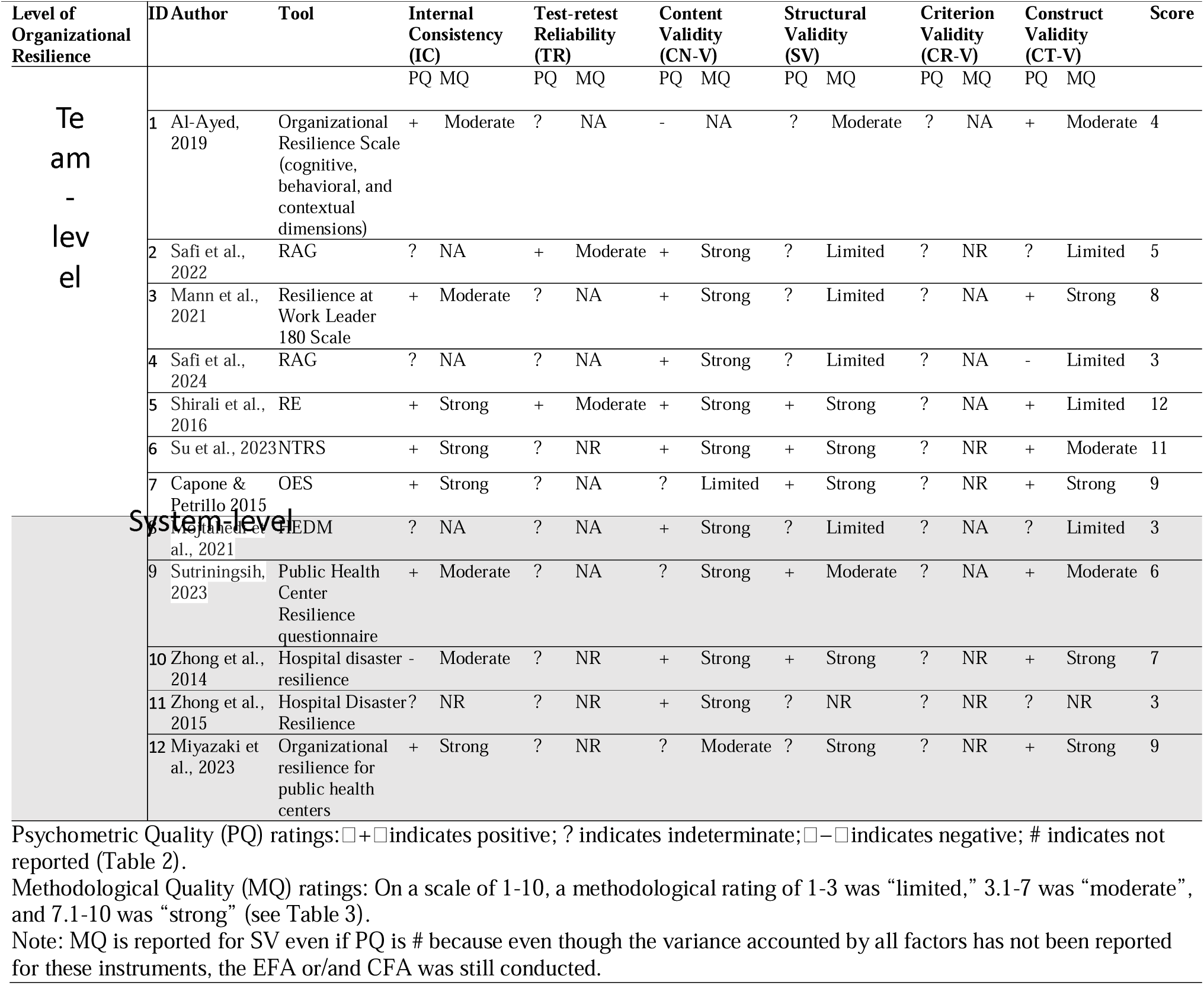
Synthesis of the quality of psychometric properties and methodological quality.

### Team-Level Organizational Resilience Instruments

Across the included studies, seven instruments assessed resilience at the team level (27–32, 37). Internal consistency was reported for five of these instruments, reflecting the questionnaire-based, self-reported nature of the measures. Strong internal consistency was observed for the NTRS (α = 0.944) (31). The Resilience Engineering (37) questionnaire demonstrated the highest internal consistency among team-level instruments (α = 0.951). Similarly, the RAG showed excellent internal consistency (α = 0.93) in healthcare team settings (30). Overall, internal consistency was acceptable to excellent, with Cronbach’s alpha being the most commonly reported indicator.

Test–retest reliability was reported in only two studies (30, 37). The Resilience Engineering questionnaire demonstrated strong temporal stability, with an intraclass correlation coefficient (ICC) of 0.95 (37). The RAG also demonstrated evidence of test–retest reliability through comparison of organizational profiles collected across two consecutive years (2021–2022) and a pilot study, indicating stability of the measured constructs over time (30).

Content validity was established for six of these team-level instruments using structured and theory-driven approaches, including expert review, Delphi methods, pilot testing, and formal calculation of item- and scale-level content validity indices. Notably, three instruments achieved the highest COSMIN ratings for content validity (30, 31, 37). The NTRS demonstrated excellent content validity following expert revision, with a scale-level content validity index (S-CVI) of 0.99 (31). The RAG similarly showed strong content validity based on Delphi consensus and expert review, with an S-CVI/Ave of 0.97 (30). The Resilience Engineering questionnaire also demonstrated excellent content validity, supported by expert review, content validity ratio (CVR; all items > 0.75), and an overall CVI of 0.85 (37).

Structural validity was evaluated in five studies using exploratory factor analysis, confirmatory factor analysis, or principal component analysis. The NTRS demonstrated a unidimensional structure with excellent model fit indices, explaining 72.33% of the total variance (31). A good model fit was also reported for the Resilience Engineering questionnaire, supporting its theoretical dimensional structure (37).

Criterion validity could not be assessed for any team-level instrument due to the absence of an established gold-standard measure of organizational or team resilience. However, construct validity was reported for all included studies and was generally supported through hypothesis testing or evidence from average variance extracted (AVE) and composite reliability (CR). For example, the NTRS demonstrated strong construct validity, with an AVE of 0.679 and a CR of 0.944 (31). Other instruments showed theoretically consistent associations with related organizational constructs.

Based on the synthesis of psychometric evidence presented in **Table 4**, the Resilience Engineering questionnaire (37) demonstrated the most comprehensive psychometric profile among team-level instruments, despite the absence of criterion validity evidence, which reflects the current lack of a gold-standard resilience measure. The NTRS (31) also achieved a high overall psychometric score; however, evidence for test–retest reliability and external construct validation remains limited, highlighting the need for further validation studies.

### System-Level Organizational Resilience Instruments

Five instruments assessed resilience at the facility or system level (33–36, 38). Internal consistency was reported for three instruments (34, 35). The Public Health Center Resilience questionnaire (34) showed strong internal consistency (Cronbach’s α = 0.84). Similarly, the Hospital Disaster Resilience Index developed by Zhong et al. (35) demonstrated acceptable reliability (α = 0.744), with most domain-level coefficients exceeding 0.60. Lastly, organizational resilience for public health centers (37) showed a high internal consistency of α =0.88.

None of the facility- or system-level instruments reported test–retest reliability or criterion validity. In contrast, content validity was addressed in all six studies and was most established through expert review, Delphi techniques, or alignment with established theoretical frameworks. Several instruments demonstrated strong evidence of content validity, including the Hospital Emergency and Disaster Management (HEDM) Index (33) and the organizational resilience for public health centers developed by Miyazaki et al. (38), which was grounded in system- and human-level resilience theory and prior qualitative research conducted by the same team.

Construct validity was reported for four of the organizational-level instruments and was generally supported through theoretical alignment rather than formal hypothesis testing. For example, the HEDM (33) demonstrated construct validity through expert-defined sub-attributes mapped to theoretical domains and operationalized using a multi-criteria decision-analysis approach (TOPSIS), providing partial support for the underlying construct. The Organizational resilience for public health centers (38) similarly, demonstrated construct validity through confirmatory factor analysis and theory-consistent relationships with related constructs.

Based on the psychometric synthesis presented in **Table 4**, the Organizational resilience for public health centers (38) demonstrated a comprehensive psychometric profile among facility- and system-level instruments, assessing structural, non-structural, and functional resilience of healthcare facilities in disaster-prone settings. the Hospital Disaster Resilience developed by Zhong et al. (35) and the Public Health Center Resilience questionnaire developed by Sutriningsih et al. (34) also showed the most promising psychometric properties. However, all three instruments have been evaluated in a limited number of studies, and further validation across diverse healthcare contexts is required to confirm their robustness and generalizability.

## Discussion

This systematic review critically evaluated and synthesized the psychometric properties and methodological quality of instruments assessing organizational resilience in health facilities. By applying stringent inclusion criteria that required explicit psychometric evaluation and a focus on collective (team, facility, and system) resilience constructs, this review identified a relatively small but methodologically coherent body of evidence. The findings highlighted both progress in the measurement of organizational resilience in healthcare and persistent gaps in conceptual clarity and psychometric rigor.

Overall, the review revealed substantial heterogeneity in how organizational resilience is conceptualized and operationalized across instruments. While several tools demonstrated acceptable to strong internal consistency and content validity, comprehensive evaluation across key psychometric domains remains uncommon. Consistent with prior methodological reviews, internal consistency was the most frequently reported property, whereas test–retest reliability and criterion validity were rarely assessed (14, 17, 39, 40). This imbalance limits confidence in the temporal stability and predictive utility of existing resilience measures.

Among the seven team-level instruments included in this review, the Resilience Engineering questionnaire (37) and the NTRS (31) demonstrated the most robust psychometric profiles. Both instruments showed strong internal consistency and supported structural validity through factor analytic methods. The Resilience Engineering questionnaire (37) further demonstrated moderate test–retest reliability, suggesting stability of the measured constructs over time. Similarly, the RAG (30, 32) showed strong content validity and evidence of temporal stability based on repeated organizational assessments, supporting its use for longitudinal resilience monitoring within healthcare teams.

Despite these strengths, none of the team-level instruments demonstrated criterion validity, reflecting the absence of a gold-standard measure for organizational or team resilience in healthcare. Although construct validity was generally supported through hypothesis testing or convergent evidence (e.g., AVE and composite reliability), external validation against objective organizational outcomes or system performance indicators remains limited. These findings underscore the need for future studies to move beyond internal validation and examine the practical and predictive relevance of team resilience instruments.

At the facility and system levels, psychometric evidence was more limited, and fewer instruments met the inclusion criteria. Among the five identified tools, Organizational resilience for public health centers by Miyazaki (38) demonstrated the most comprehensive psychometric profile, with acceptable internal consistency and evidence supporting its structural validity. The hospital disaster resilience developed by Zhong et al. (35) also showed acceptable reliability and theoretically grounded construct validity related to disaster preparedness, infrastructure safety, and emergency response capacity.

The Public Health Center Resilience questionnaire (34) contributed to this group by explicitly integrating system-level and human resilience dimensions and demonstrating promising construct validity through factor analysis and theory-consistent associations. However, similar to other facility-level instruments, evidence for test–retest reliability and criterion validity was absent. This pattern reflects broader methodological challenges in measuring organizational resilience at the system level, where objective benchmarks and standardized validation approaches remain underdeveloped.

A central finding of this review was the uneven application of psychometric standards across organizational resilience instruments. While content validity was commonly addressed, often through expert review, Delphi methods, or theoretical alignment, other essential properties, including temporal stability and criterion-related validity, were rarely evaluated. Many instruments appear to be in early stages of development, limiting their suitability for benchmarking, comparative analysis, or evaluation of resilience-building interventions.

The limited number of eligible instruments identified in this review does not indicate a lack of conceptual interest in organizational resilience, but rather highlights a gap between conceptual frameworks and rigorously validated measurement tools. Numerous studies propose resilience indicators or assessment frameworks; however, relatively few translate these concepts into psychometrically tested instruments suitable for empirical application in healthcare settings.

Moreover, the COSMIN checklist underscored the methodological interdependence between structural validity and internal consistency, as factor analysis should precede reliability testing to ensure the coherence of scale items (17, 40). Despite this, several instruments reported alpha coefficients without supporting factor structures, highlighting concerns raised in psychometric best practices (13, 14).

Additionally, one of the central challenges encountered in this review was the lack of conceptual uniformity across studies regarding what constitutes “organizational resilience.” This variation affects not only instrument development but also interpretation and applicability. While some instruments aimed to measure resilience at the level of entire health systems, others focused on facility-level capacities or subcomponents such as workforce responsiveness or leadership. As a result, our review may reflect some subjective bias in the inclusion and scoring of instruments, particularly in borderline cases. We attempted to mitigate this by relying on predefined inclusion criteria and consensus-based decisions among reviewers.

Consistent with Ignatowicz et al. (8), this review also identified the absence of intersectional approaches in resilience measurement. Resilience, as a multifaceted and context-dependent phenomenon, intersects with multiple structural determinants, including socio-economic status and organizational power dynamics (41). Future research should integrate intersectional and systems-thinking approaches into the development and validation of resilience measures to capture the complex realities of diverse healthcare workforces and facilities (42). Integrative frameworks are needed to bridge this gap, ensuring resilience is measured across nested levels of influence, from frontline care delivery to systemic preparedness and adaptability.

### Limitations

By focusing exclusively on instruments with reported psychometric evaluation and distinguishing between team-level and system-level measurement, this review provides a focused and methodologically rigorous synthesis of the current evidence base. The findings offer practical guidance for researchers and health system leaders seeking validated tools to assess organizational resilience.

However, this review had several limitations. First, while two researchers conducted independent data extraction, intercoder reliability was not formally reported. Second, our review also focused on peer-reviewed articles published in English and psychometrically validated measures that are published in indexed databases and meet standard psychometric reporting, which may have excluded gray literature and unpublished tools developed for programmatic use. Additionally, the high proportion of conference presentations that could not be retrieved and eventually excluded from the study reflects a lack of robust empirical studies in this area.

Third, the absence of a universal definition of organizational resilience in healthcare may have introduced subjectivity in determining the eligibility of instruments. Although we relied on consensus and existing frameworks, variability in conceptualization and scope across studies remains a limitation. Further, while the review assessed six core psychometric properties, measurement invariance and cross-cultural validation were not evaluated systematically, given the focus on instrument validation at a single point in time. Additionally, the scoring system weighed all psychometric properties equally, which may not reflect their contextual relevance in different research or practice settings.

In addition, the review focused primarily on psychometric properties assessed through classical test theory, as operationalized through the COSMIN checklist. While this approach remains widely used, it may not capture insights from modern psychometric frameworks such as item response theory, Rasch modeling (e.g., one-dimensionality diagnostics), or advanced confirmatory factor analysis and structural equation modeling techniques, which are increasingly applied in the evaluation of resilience scales. Future reviews could incorporate these methods to provide a more comprehensive evaluation of scale dimensionality and performance.

Finally, while the COSMIN checklist provided a valuable framework for assessing methodological quality, it was originally developed for PROMs and may not fully account for the complexities and contextual nuances of structural-level tools. Although we adapted its criteria for organizational measures, the comparability of COSMIN scores across fundamentally different types of instruments (e.g., team vs. system-level tools) should be interpreted with caution. Moreover, the absence of a gold standard tool for appraising the methodological quality of system-level instruments introduces subjectivity into our ratings. We addressed this by involving multiple independent raters, reconciling discrepancies, and avoiding direct one-to-one score comparisons across divergent instrument types.

### Future Directions

Future research should prioritize the continued development and rigorous psychometric validation of instruments assessing organizational resilience at team and facility/system levels in healthcare settings. In particular, greater emphasis is needed on evaluating underreported measurement properties, including test–retest reliability, criterion validity, and predictive validity, to establish the temporal stability and practical relevance of existing tools. Longitudinal validation studies are essential to determine whether resilience instruments are sensitive to change over time and capable of capturing the effects of organizational interventions or system disruptions.

Further research should also aim to strengthen construct validation by examining associations between resilience measures and objective organizational outcomes, such as safety performance, service continuity, or adaptive capacity during crises. The absence of an accepted gold-standard measure of organizational resilience remains a major barrier to criterion validation, underscoring the need for consensus-building efforts and outcome-linked benchmarking strategies in this field.

Cross-context and cross-cultural validation of organizational resilience instruments is another critical priority. Although several tools included in this review were developed or applied in diverse healthcare contexts, few studies employed advanced psychometric techniques such as measurement invariance testing or item response theory to assess the comparability of scores across settings, professional groups, or health systems. Addressing this gap will be essential to improve the generalizability and global applicability of resilience measures.

Finally, future instrument development should more explicitly capture the structural, operational, and contextual dimensions of healthcare resilience. This review identified a scarcity of instruments designed to assess structural-level resilience, including infrastructure robustness, resource flexibility, and system-level interdependencies. Given that existing methodological frameworks, such as COSMIN, were originally developed for patient-reported outcomes, there is also a need for complementary evaluation approaches tailored to organizational and system-level constructs. Advancing such frameworks will be critical for supporting robust, theory-driven measurement of resilience and for translating resilience assessment into actionable decision-making within healthcare systems.

## Conclusions

This comprehensive systematic review critically appraised the psychometric properties and methodological quality of instruments designed to measure organizational resilience among health facilities. Based on combined psychometric ratings and methodological quality scores, at the team level, the Resilience Engineering questionnaire and at the facility level, the Organizational resilience for public health centers emerged as the most psychometrically robust instruments in this review. The overall evaluation revealed that while an increasing number of instruments are being developed and adapted, only a limited subset has undergone comprehensive psychometric evaluation, particularly at the organizational and system levels. Our findings confirm the urgent need for the development and validation of psychometrically robust, culturally sensitive, and context-specific organizational resilience instruments. Additionally, a comprehensive psychometric evaluation, including test-retest reliability, criterion validity, and measurement invariance testing, remains essential to strengthen the evidence base for these instruments. Given the limited availability of system-level resilience measures with adequate psychometric support, future research should also prioritize the development of new tools that comprehensively address organizational resilience within diverse health settings.

## Data Availability

All data produced in the present work are contained in the manuscript.

## Acknowledgments

### Funding

The study was funded by NIH/NIAID (Grant #R01AI174892).

### Author Contribution

**Atena Pasha** (Investigation, Methodology, Project administration, Resources, Formal analysis, Writing – original draft, Writing – review and editing), **Kaylin Morris, Karolina Kaczmarski** (Investigation, Data curation, Formal analysis), **Xiaoming Li** (Conceptualization, Supervision, Writing – review and editing), **Shan Qiao** (Conceptualization, Supervision, Methodology, Writing – review and editing).

All authors read and approved the final manuscript.

### Ethics approval and consent to participate

Not applicable.

### Consent for publication

Not applicable.

### Availability of data and materials

No data was generated by this study. All data and materials, including data extracted from included studies, data used for all analyses, and any other materials used in the review, are available in this manuscript and its supplementary materials.

