## Supplemental document for "Measurement Instruments Assessing Organizational Resilience in Health Facilities: A Systematic Review of Psychometric Properties"

**Appendix 1.** Summary of search results.

| **Database** | **Search String** |
| --- | --- |
| **PsycINFO (EBSCOhost)** | ( TI ( (measure* N8 resilien*) OR (instrument* N8 resilien*) OR (scale* N8 resilien*) OR (tool* N8 resilien*) OR (test* N8 resilien*) OR (questionnaire* N8 resilien*) OR (survey* N8 resilien*) OR (assess* N8 resilien*) OR “organizational resilien*” OR “organisational resilien*” ) OR AB ( (measure* N8 resilien*) OR (instrument* N8 resilien*) OR (scale* N8 resilien*) OR (tool* N8 resilien*) OR (test* N8 resilien*) OR (questionnaire* N8 resilien*) OR (survey* N8 resilien*) OR (assess* N8 resilien*) OR “organizational resilien*” OR “organisational resilien*” ) ) AND ( ( DE "Hospitals" OR DE "Clinics" ) OR TI ( “health system*” OR “health care system*” OR “health institution*” OR “health organization*” OR “health organisation*” OR “delivery of health care” OR hospital* OR “health facilit*” OR “health care facilit*” OR “health service*” OR “healthcare setting*” OR “health delivery” OR “health center*” OR clinic OR clinics OR “healthcare sector*” OR “health care sector*” OR “health center*” ) OR AB ( “health system*” OR “health care system*” OR “health institution*” OR “health organization*” OR “health organisation*” OR “delivery of health care” OR hospital* OR “health facilit*” OR “health care facilit*” OR “health service*” OR “healthcare setting*” OR “health delivery” OR “health center*” OR clinic OR clinics OR “healthcare sector*” OR “health care sector*” OR “health center*” ) ) AND LA English NOT PT Reviews |
| **MEDLINE (PubMed)** | ("measure resilience"[tiab:~8] OR "measure resiliency"[tiab:~8] OR "instrument resilience"[tiab:~8] OR "instrument resiliency"[tiab:~8] OR "scale resilience"[tiab:~8] OR "scale resiliency"[tiab:~8] OR "tool resilience"[tiab:~8] OR "tool resiliency"[tiab:~8] OR "measurement resilience"[tiab:~8] OR "measurement resiliency"[tiab:~8] OR "test resilience"[tiab:~8] OR "test resiliency"[tiab:~8] OR "questionnaire resilience"[tiab:~8] OR "questionnaire resiliency"[tiab:~8] OR "survey resilience"[tiab:~8] OR "survey resiliency"[tiab:~8] OR "assessment resilience"[tiab:~8] OR "assessment resiliency"[tiab:~8] OR "assess resilience"[tiab:~8] OR "assess resiliency"[tiab:~8] OR "organizational resilien*"[tiab] OR "organisational resilien*"[tiab]) AND ("Delivery of Health Care"[Mesh:NoExp] OR "Hospitals"[Mesh:NoExp] OR "Health Facilities"[Mesh:NoExp] OR "Health Services"[Mesh] OR "Health Care Sector"[Mesh] OR "health system*"[tiab] OR "health care system*"[tiab] OR "health institution*"[tiab] OR "health organization*"[tiab] OR "health organisation*"[tiab] OR "delivery of health care"[tiab] OR hospital*[tiab] OR "health facilit*"[tiab] OR "health care facilit*"[tiab] OR "health service*"[tiab] OR "healthcare setting*"[tiab] OR "health delivery"[tiab] OR "health center*"[tiab] OR clinic[tiab] OR clinics[tiab] OR "healthcare sector*"[tiab] OR "health care sector*"[tiab]) AND English[la] NOT ("Editorial"[pt] OR "Comment"[pt] OR "Review"[pt] OR "Systematic Review"[pt] OR "Meta-Analysis"[pt]) |
| **Embase (Elsevier)** | ('organizational resilience'/de OR ((measure* NEAR/8 resilien*):ab,ti) OR ((instrument* NEAR/8 resilien*):ab,ti) OR ((scale* NEAR/8 resilien*):ab,ti) OR ((tool* NEAR/8 resilien*):ab,ti) OR ((test* NEAR/8 resilien*):ab,ti) OR ((questionnaire* NEAR/8 resilien*):ab,ti) OR ((survey* NEAR/8 resilien*):ab,ti) OR ((assess* NEAR/8 resilien*):ab,ti) OR 'organizational resilien*':ab,ti OR 'organisational resilien*':ab,ti) AND ('health care'/de OR 'health care organization'/de OR 'health care delivery'/de OR 'hospital'/de OR 'health care facility'/de OR 'health service'/de OR 'health center'/exp OR 'health system*':ab,ti OR 'health care system*':ab,ti OR 'health institution*':ab,ti OR 'health organization*':ab,ti OR 'health organisation*':ab,ti OR 'delivery of health care':ab,ti OR hospital*:ab,ti OR 'health facilit*':ab,ti OR 'health care facilit*':ab,ti OR 'health service*':ab,ti OR 'healthcare setting*':ab,ti OR 'health delivery':ab,ti OR clinic:ab,ti OR clinics:ab,ti OR 'healthcare sector*':ab,ti OR 'health care sector*':ab,ti OR 'health center*':ab,ti) AND [english]/lim NOT 'review'/it |
| **Web of Science (Clarivate)** | ((TI=((measure* NEAR/8 resilien*) OR (instrument* NEAR/8 resilien*) OR (scale* NEAR/8 resilien*) OR (tool* NEAR/8 resilien*) OR (test* NEAR/8 resilien*) OR (questionnaire* NEAR/8 resilien*) OR (survey* NEAR/8 resilien*) OR (assess* NEAR/8 resilien*) OR “organizational resilien*” OR “organisational resilien*”)) OR AB=((measure* NEAR/8 resilien*) OR (instrument* NEAR/8 resilien*) OR (scale* NEAR/8 resilien*) OR (tool* NEAR/8 resilien*) OR (test* NEAR/8 resilien*) OR (questionnaire* NEAR/8 resilien*) OR (survey* NEAR/8 resilien*) OR (assess* NEAR/8 resilien*) OR “organizational resilien*” OR “organisational resilien*”)) AND ((TI=(“health system*” OR “health care system*” OR “health institution*” OR “health organization*” OR “health organisation*” OR “delivery of health care” OR hospital* OR “health facilit*” OR “health care facilit*” OR “health service*” OR “healthcare setting*” OR “health delivery” OR “health center*” OR clinic OR clinics OR “healthcare sector*” OR “health care sector*” OR “health center*” )) OR AB=(“health system*” OR “health care system*” OR “health institution*” OR “health organization*” OR “health organisation*” OR “delivery of health care” OR hospital* OR “health facilit*” OR “health care facilit*” OR “health service*” OR “healthcare setting*” OR “health delivery” OR “health center*” OR clinic OR clinics OR “healthcare sector*” OR “health care sector*” OR “health center*” )) |
| **CINAHL (EBSCOhost)** | ( TI ( (measure* N8 resilien*) OR (instrument* N8 resilien*) OR (scale* N8 resilien*) OR (tool* N8 resilien*) OR (test* N8 resilien*) OR (questionnaire* N8 resilien*) OR (survey* N8 resilien*) OR (assess* N8 resilien*) OR “organizational resilien*” OR “organisational resilien*” ) OR AB ( (measure* N8 resilien*) OR (instrument* N8 resilien*) OR (scale* N8 resilien*) OR (tool* N8 resilien*) OR (test* N8 resilien*) OR (questionnaire* N8 resilien*) OR (survey* N8 resilien*) OR (assess* N8 resilien*) OR “organizational resilien*” OR “organisational resilien*” ) ) AND ( ( (MH "Health Care Delivery") OR (MH "Hospitals") OR (MH "Health Facilities") OR (MH "Health Services") ) OR TI ( “health system*” OR “health care system*” OR “health institution*” OR “health organization*” OR “health organisation*” OR “delivery of health care” OR hospital* OR “health facilit*” OR “health care facilit*” OR “health service*” OR “healthcare setting*” OR “health delivery” OR “health center*” OR clinic OR clinics OR “healthcare sector*” OR “health care sector*” OR “health center*” ) OR AB ( “health system*” OR “health care system*” OR “health institution*” OR “health organization*” OR “health organisation*” OR “delivery of health care” OR hospital* OR “health facilit*” OR “health care facilit*” OR “health service*” OR “healthcare setting*” OR “health delivery” OR “health center*” OR clinic OR clinics OR “healthcare sector*” OR “health care sector*” OR “health center*” ) ) AND LA English NOT PT Reviews |

**Appendix 2.** Quality criteria used to assess psychometric properties of measures (Terwee et al., 2007; Terwee et al., 2012).

| Psychometric property | Definition | Rating | Quality criteria |
| --- | --- | --- | --- |
| Internal consistency | The extent to which the items correlate, indicating that the overall instrument is measuring the same construct | + | Data from adequate sample used to conduct factor analysis and Cronbach’s α > 0.70 |
|  |  | ? | Cronbach’s α not reported |
|  |  | − | Cronbach’s α < 0.70 on any one factor |
| Test-retest Reliability | The degree to which the scores are free from measurement error | + | Significant ICC/weighted kappa ≥ 0.70 or significant Pearson’s correlation for all factors |
|  |  | ? | ICC/weighted kappa or Pearson’s correlation not reported |
|  |  | − | ICC/weighted kappa < 0.70 or Pearson’s correlation insignificant for at least one factor |
| Content validity | The extent to which the items reflect the construct being assessed | + | Extensive literature review, item selection with experts, and involvement of target population (qualitative surveys, cognitive interviewing) |
|  |  | ? | Only literature review conducted but target population not involved |
|  |  | − | No information on literature review, expert panel, and involvement of target population |
| Structural validity | The degree to which the scores of the instrument adequately reflect the dimensions of the construct being assessed | + | Factor analysis demonstrates that combined set of factors explain ≥ 50% of total variance in the model |
|  |  | ? | Proportion of variance explained not reported |
|  |  | − | <50% of the total variance explained by model |
| Criterion validity | The extent to which scores on a particular questionnaire relate to a gold standard | + | More than half the correlations with “gold” standards are significant |
|  |  | ? | Criterion validity not reported |
|  |  | − | Less than half the correlations with “gold” standards are not significant |
| Construct validity | The extent to which scores on a particular questionnaire relate to other measures in a manner that is consistent with theoretically derived hypotheses | + | At least 75% of the results are in accordance with hypotheses |
|  |  | ? | Construct validity not reported |
|  |  | − | Less than 75% of hypotheses were confirmed. |
| Psychometric property ratings: + indicates positive; ? indicates indeterminate; − indicates negative  ICC Intraclass correlation coefficient | | | |

**Appendix 3.** The scoring system used to compare overall properties of the instruments*.*

| Psychometric Quality Rating | Methodological Quality Rating | Scores Assigned |
| --- | --- | --- |
| + | Strong | +3 |
| + | Moderate | +2 |
| + | Limited | +1 |
| - | Strong | -1 |
| - | Moderate | -2 |
| - | Limited | -3 |

Psychometric Quality (PQ) ratings: + indicates positive; ? indicates indeterminate; − indicates negative; # indicates not reported.

Methodological Quality (MQ) ratings: On a scale of 1-10, a methodological rating of 1-3 was “limited,” 3.1-7 was “moderate”, and 7.1-10 was “strong”.
