## Supplementary material for "Measurement Instruments Assessing Organizational Resilience in Health Facilities: A Systematic Review of Psychometric Properties": Figure

**Fig 1.** Flow diagram of the literature search and articles selection (adapted from PRISMA 2020 guidelines for systematic reviews) (23).

**Identification of studies via databases and registers**

Records identified from*:

MEDLINE (PubMed) (n = 1603)

Web of Science (Clarivate) (n = 1976)

PsycINFO (EBSCOhost) (n = 760)

CINAHL (EBSCOhost) (n = 891)

Embase (Elsevier) (n = 2249)

Records removed *before screening*:

Duplicate records removed (n = 2333)

**Identification**

Records excluded*

(n = 3764)

Records screened

(n = 5146)

Reports not retrieved**

(n = 935)

**Screening**

Reports sought for retrieval

(n = 1382)

Reports excluded:

Duplicates (n = 22)

No relevant information (n = 154)

Wrong population

(n = 47)

Wrong study design

(n = 203)

Qualitative study only (n = 7)

Reports assessed for eligibility

(n = 447)

Studies included in review

(n = 12)

**Included**

* 3,764 studies were excluded for reasons such as irrelevance to organizational resilience, lack of psychometric data, use of modeling tools without validation, or instruments focused solely on individual-level resilience constructs without linking to organizational or team-level dynamics.

** 935 full texts could not be retrieved due to access restrictions, broken links, or conference presentations without full data.
